# Estimating waning vaccine effectiveness from population-level surveillance data in multi-variant epidemics

**DOI:** 10.1101/2022.07.14.22277647

**Authors:** Hiroaki Murayama, Akira Endo, Shouto Yonekura

**Affiliations:** School of Medicine, International University of Health and Welfare, Narita, Japan; Graduate School of Social Sciences, Chiba University, Chiba, Japan; Department of Infectious Disease Epidemiology, London School of Hygiene and Tropical Medicine, London, United Kingdom; Centre for the Mathematical Modelling of Infectious Diseases, London School of Hygiene and Tropical Medicine, London, United Kingdom; School of Tropical Medicine and Global Health, Nagasaki University, Nagasaki, Japan

## Abstract

Monitoring time-varying vaccine effectiveness (e.g., due to waning of immunity and the emergence of novel variants) provides crucial information for outbreak control. Existing studies of time-varying vaccine effectiveness have used individual-level data, most importantly dates of vaccination and variant classification, which are often not available in a timely manner or from a wide range of population groups. We present a novel Bayesian framework for estimating the waning of variant-specific vaccine effectiveness in the presence of multi-variant circulation from population-level surveillance data. Applications to simulated outbreak and COVID-19 epidemic in Japan are also presented. Our results show that variant-specific waning vaccine effectiveness estimated from population-level surveillance data could approximately reproduce the estimates from previous test-negative design studies, allowing for rapid, if crude, assessment of the epidemic situation before fine-scale studies are made available.

**Author summary:** The emergence of immunity-escaping SARS-CoV-2 variants and the waning of vaccine effectiveness have highlighted the need for near-real-time monitoring of variant-specific protection in the population to guide control efforts. However, standard epidemiological studies to this end typically require access to detailed individual-level dataset, which may not be timely available in an ongoing outbreak. A more convenient and less resource-intensive approach using routinely-collected data could complement such studies by providing tentative estimates of waning vaccine effectiveness until the conclusive evidence becomes available. In this paper, we propose a novel Bayesian framework for estimating waning vaccine effectiveness against multiple co-circulating variants that requires only population-level surveillance data. Using simulated outbreak data of multiple variants,we showed that the proposed method can plausibly recover the ground truth from population-level data. We also applied the proposed method to empirical COVID-19 data in Japan, which yielded estimates that are overall in line with those derived from studies using individual-level data.

## Introduction

Rapid development and distribution of highly effective vaccines against severe acute respiratory syndrome coronavirus 2 (SARS-CoV-2) [1], with the earliest mass vaccination campaign initiated in December 2020 [2], was initially expected to swiftly end the coronavirus disease 2019 (COVID-19) pandemic. However, the waning of vaccine-induced protection observed among those who are past months since vaccination [3, 4], along with the emergence of novel variants that exhibit higher transmission potentials and/or an immune escaping features [5], has compromised this expectation, and the long-term outlook of the outbreak control including vaccine booster strategies has become increasingly complicated [6, 7]. As of June 2022, many countries have provided the third (i.e., booster) doses of COVID-19 vaccines to the general population who are past a specified period of time since the second dose [2]; moreover, some countries have been offering the fourth doses to some high-risk groups [8]. With dynamically-changing population immunity landscapes in the presence of emerging variants, monitoring the strength and duration of vaccine-induced protection has become critically important in informing such dosing schedules and decision making on supplemental control measures.

A number of studies have assessed the waning of vaccine-induced protection by estimating the vaccine effectiveness as a function of time since vaccination [3, 9–15]. Some of those studies reported variant-specific waning effects, which suggested that the protection against the Omicron variant wanes quickly than the Delta variant [9, 14]. While these studies have provided fine-scale and robust findings on the waning of vaccine effectiveness, they are not necessarily the most rapidly/easily implemented studies as they essentially require detailed individual-level data, which at least should include timing of infection, the most-recent date of vaccination, and (where variant-specific waning is of interest) either sequence data or proxy of variant identification (e.g., S-gene target failure [16]). As a result, those data were often not available in the most timely manner or only available in limited (resource-rich) settings. Given the continuously evolving situations of the SARS-CoV-2 circulation, e.g., as with the recent emergence of the Omicron sublineages [17], a more convenient and less resource-intensive, if crude, approach to monitoring time-varying vaccine effectiveness would be warranted, which could provide tentative estimates until fine-scale studies eventually become available.

In this study, we propose a novel Bayesian framework for estimating waning vaccine effectiveness from population-level surveillance data in the presence of multi-variant circulation. Assuming either parametric or semi-parametric temporal patterns of waning, our model allows for estimation of variant-specific time-varying vaccine effectiveness. As application examples, we used our method to estimate the waning vaccine effectiveness in simulated and real-world outbreak datasets.

## Materials and methods

### Modelling framework

We construct a discrete renewal process that describes the incidence of new infections among the unvaccinated and vaccinated populations and in week *t*. As partially vaccinated individuals (i.e., those with only one dose of COVID-19 vaccines with a two-dose regimen) typically account for only small fraction of the population over time, hereafter we assume that individuals obtain “vaccinated” status only when they are fully vaccinated according to the regimen. Let *i*_*t*_ and *j*_*t*_ be the number of new infections among the unvaccinated and vaccinated populations in week *t*, respectively. Assuming proportionate mixing [18–20], the renewal process among unvaccinated and vaccinated populations can be modelled as

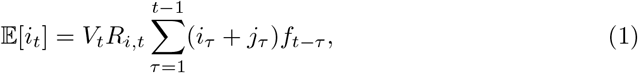

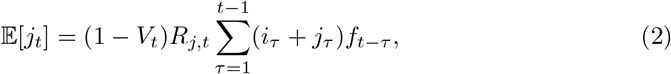

where 𝔼[·] denotes the operator of expectation, *R*_*i,t*_ is the mean number of unvaccinated secondary cases generated by a primary case in a wholly unvaccinated population in week *t*, and *R*_*j,t*_ is the mean number of secondary transmissions in a wholly vaccinated population in week *t*, and *f*_*t*_ is the probability mass function for the generation time (i.e., time interval between infections of primary and secondary case pairs). *V*_*t*_ represents the cumulative proportion vaccinated by week *t*.

*R*_*i,t*_ and *R*_*j,t*_ can be related using the variant-weighted cross sectional protection (VCP; denoted as *ϵ*_*t*_):

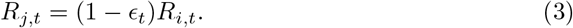

Here we define the VCP as the population-level relative risk reduction in week *t*, weighted by the relative frequency of the circulating variants. VCP can be computed from the variant-specific waning curves of vaccine effectiveness and the time-varying relative frequency of variants:

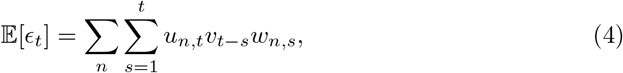

where *u*_*n,t*_ is the relative frequency of variant *n* in week *t, v*_*n,t*_ is the weekly rate of vaccination per capita, and *w*_*n,s*_ represents waning vaccine effectiveness against variant *n* as a function of the number of weeks since vaccination *s*.

We model the functional form of waning vaccine effectiveness *w*_*n,s*_ in two approaches, i.e., (i) parametric and (ii) semi-parametric approaches.

In the parametric approach, we assume an exponential decay of vaccine effectiveness:

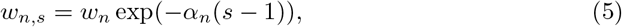

where *w*_*n*_(≥ 0) and *α*_*n*_(≥ 0) denote the initial effectiveness and the weekly decay constant, respectively. Alternatively, a logistic curve is used as part of the sensitivity analysis, given as

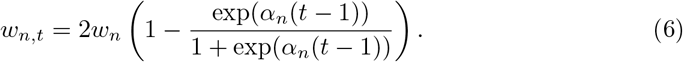

In the semi-parametric approach, we use a natural cubic spline on the logit of *w*_*n,s*_, i.e.,

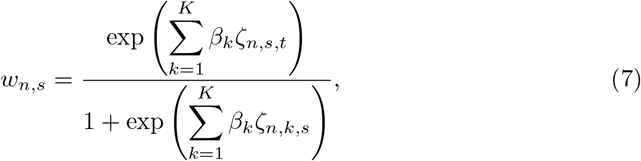

where *ζ*_*n,k,t*_ (*k* = 1, …, *K*) are natural cubic spline basis functions with *K* equally spaced internal knots, and *β*_*k*_ is the spline coefficient [21]. We denote the values at the spline knots by *y*_*n,k*_, which we treat as free parameters in the analysis.

### Bayesian inference

To account for the individual-level heterogeneity in vaccine-induced protection, we introduce an overdispersion in VCP via hierarchical modelling. As VCP is defined as an average over those exposed in week *t*, its variance is expected to be inversely proportional to the sample size (i.e., the number of exposed vaccinated individuals in week *t*), which we assume to be proportional to *j*_*t*_. As such, *ϵ*_*t*_ is assumed to be beta-distributed with a mean 𝔼[*ϵ*_*t*_]:

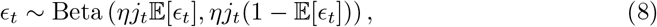

where *η* is a hyperparamter that controls the variance of *ϵ*_*t*_. For *ηj*_*t*_ ≫ 1, we get 𝕍 [*ϵ*_*t*_] ≈𝔼 [*ϵ*_*t*_](1 −𝔼 [*ϵ*_*t*_])*/ηj*_*t*_.

We then construct the Poisson likelihood for observed incidence as

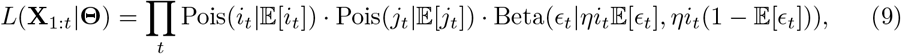

where **X**_1:*t*_ is the time-series of observed data: *i*_*t*_, *j*_*t*_, *u*_*t*_, and *v*_*t*_. **Θ** is the set of unknown parameters: (*w*_*n*_, *α*_*n*_, *η, R*_*i,t*_) in the parametric approach; and (*y*_*n,k*_, *η, R*_*i,t*_) in the semi-parametric approach. The parameters and corresponding prior distributions are summarised in Table 1.

**Table 1.** Model parameters and corresponding priors.

| Parameter | Description | Prior |
| --- | --- | --- |
| $R_{i,t}$ | Mean secondary transmissions in a wholly unvaccinated population ($R_{i,t} \geq 0$) | LeftTruncatedNormal(1, 2) |
| $w_n$ | Initial vaccine effectiveness ($w_n \geq 0$) | Uniform(0, 2) |
| $\alpha_n$ | Exponential decay constant ($\alpha_n \geq 0$) | HalfNormal(10) |
| $\text{inv\_logit}(y_{n,k})$ | Vaccine effectiveness against variant $n$ at the spline knots | Beta(5, 2) |
| $\eta$ | Hyperparameter controlling the variance of $\epsilon_t$ ($\eta \geq 0$) | HalfNormal(100) |

### Application examples

We applied our method to a simulated outbreak dataset and COVID-19 dataset in Japan to estimate waning vaccine effectiveness in the presence of multi-variant circulation.

### Simulated outbreak

We first used a simulated outbreak dataset of four variants (A, B, C, and D with different transmission potentials) to assess if our model can recover the original parameters from the generated outbreak data. The distribution of generation time *f*_*t*_ was borrowed from that of serial interval for COVID-19 [22]. The variant-specific basic reproduction numbers were denoted as *λ*_*n*_ = *r*_*n*|*A*_*λ*_*A*_, where *r*_*n*|*A*_ represents the relative transmissibility of variant *n* to variant A. The effective reproduction number for variant *n* is then given as *R*_*n,i,t*_ = (1 − *ρ*_*t*_)Λ_*n,t*_, where *ρ*_*t*_ represents the time-varying intensity of non-pharmaceutical interventions (NPIs). We assumed that the waning of vaccine effectiveness follows a logistic curve in Eq (6). Cross-sectional protection from vaccines against each variant is modelled as

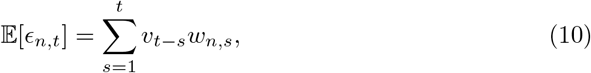

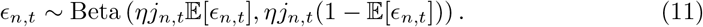

The initial numbers of cases of variant A, *i*_*A,t*_ and *j*_*A,t*_, were set as (*i*_1_, *i*_2_) = (5300, 5200) and (*j*_1_, *j*_2_) = (3, 5). Those of other variants were (*i*_*B*,15_, *i*_*C*,40_, *i*_*D*,55_) = (30, 30, 5) and (*j*_*B*,15_, *j*_*C*,40_, *j*_*D*,55_) = (15, 30, 15), respectively. We assumed that the number of secondary transmissions of variant *n* in a wholly unvaccinated population by week (Λ_*n,t*_) randomly fluctuates around the mean transmission potential (*λ*_*n,t*_):

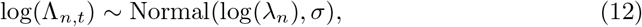

where *σ* denotes the standard deviation of the normal distribution. The parameter specifications for the simulation can be found in Table 2.

**Table 2.** Summary of model parameters for the simulation.

| Parameter | Description | Selected value |
| --- | --- | --- |
| $\lambda_A$ | Mean secondary transmissions of variant A per primary case to unvaccinated individuals under no NPIs implemented | 1.3 |
| $r_{B\vert A}$ | Relative transmissibility of variant B to A | 1.85 |
| $r_{C\vert B}$ | Relative transmissibility of variant C to B | 2.1 |
| $r_{D\vert C}$ | Relative transmissibility of variant D to C | 3 |
| $\rho_t$ | Effectiveness of NPIs on curtailing infection | 0 ($t=1, \dots, 29$)<br>0.5 ($t=30, \dots, 54$)<br>0.85 ($t=55, \dots, 73$) |
| $w_A$ | Initial vaccine effectiveness against variant A | 0.95 |
| $w_B$ | Initial vaccine effectiveness against variant B | 0.90 |
| $w_C$ | Initial vaccine effectiveness against variant C | 0.90 |
| $w_D$ | Initial vaccine effectiveness against variant D | 0.80 |
| $\alpha_A$ | Exponential decay constant of variant A | 0.01 |
| $\alpha_B$ | Exponential decay constant of variant B | 0.02 |
| $\alpha_C$ | Exponential decay constant of variant C | 0.01 |
| $\alpha_D$ | Exponential decay constant of variant D | 0.03 |
| $\eta$ | Hyperparameter controlling the variance of $\epsilon_{n,t}$ ($n=A, B, C, D$) | 0.2 |
| $\sigma$ | Standard deviation of the normal distribution for the daily random fluctuation of the mean transmissibility | 0.1 |

### COVID-19 in Japan (2021–2022)

We also applied our method to the COVID-19 outbreak in Japan from the week of 27 September 2021 to the week of 22 March 2022. The end date of the study period was chosen to exclude the possible impact of booster doses (because booster status was not reported in the dataset). Case counts by vaccination status and the daily number of newly vaccinated individuals were publicly available [23–25]. A total 3,416,586 cases were included. Unvaccinated cases under the age of 11 were excluded because the administration of vaccine to this population had not been approved during the timeframe. The weekly proportion of variants was obtained from the figures in [26].

We compared the estimated vaccine effectiveness in our model with the estimates from the previous test-negative case controls studies (in Canada against Delta and in Japan against Omicron) [13, 14].

### Markov chain Monte Carlo

We estimated the parameters for each of the datasets using the No-U-Turn-Sampler algorithm. For the analysis of simulated outbreak data, 6,000 Markov chain Monte Carlo (MCMC) samples were obtained from two chains, where the first 500 samples of each chain were discarded as warm-up. For the analysis of COVID-19 epidemic in Japan, we obtained 10,000 samples from two chains after discarding the first 500 samples as warm-up. The results of MCMC sampling showed an R-hat statistic of below 1.02 and an effective sample size of at least 400. All the analysis was conducted with the ‘*rstan*’ package in R version 4.0.2.

## Results

### Application example using simulated outbreak data

We simulated an outbreak consisting of four variants replacing one another over time (Fig 2). Vaccine roll-out and waning of variant-specific vaccine effectiveness were also simulated along the outbreak progression. For each variant *n*, The unvaccinated and vaccinated cases were simulated by applying the renewal process in Eq (1) and Eq (2) to each variant (Fig 2 (B)). We then estimated the waning vaccine effectiveness against each of the variants from the simulated data. The results suggested that our parametric approach with an exponential decay assumption plausibly recovered the ground truth vaccine effectiveness (Fig 3 (A)–(D)). The comparison between the proportion vaccinated and VCP shows that the model captured a gradual decline of VCP due to waning, in particular after the replacement by variant D, against which vaccines were assumed to be less effective and to wane more rapidly (Fig 3 (E)). Meanwhile, the semi-parametric approach yielded overall consistent yet slightly unstable/biased estimates, especially around the tail part of the waning curves (Fig 4).

**Fig 1.**
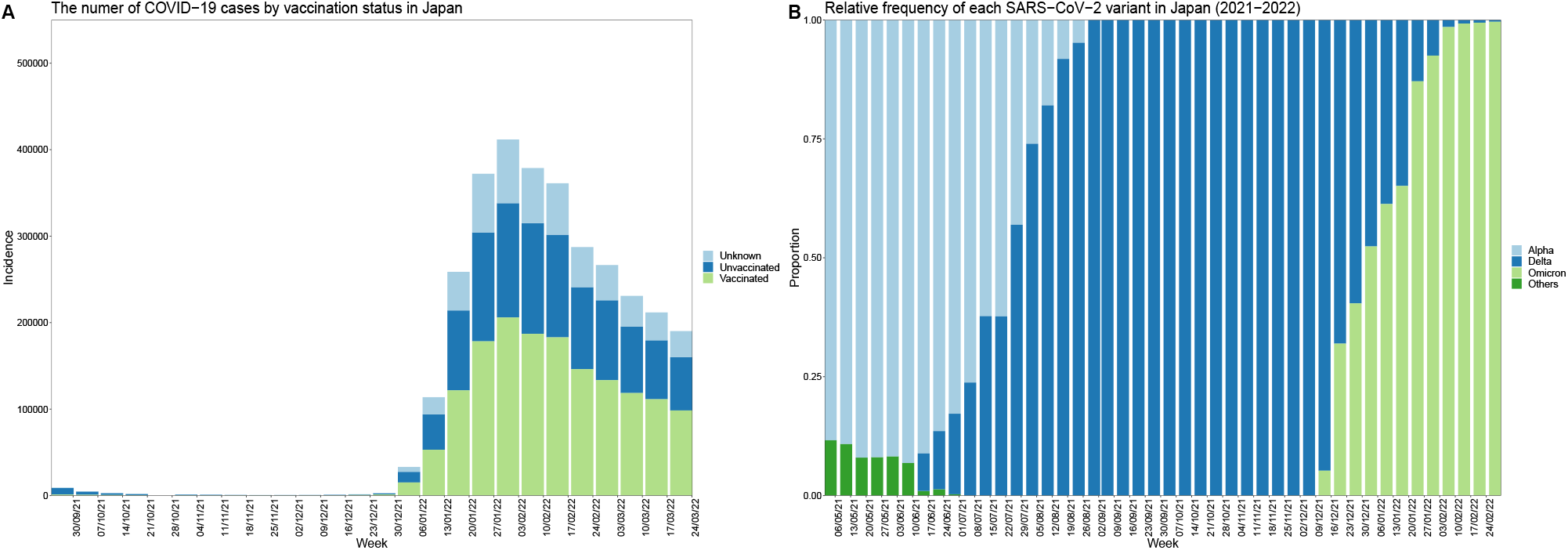
Reported weekly number of COVID-19 cases with vaccine status and proportion of each SARS-CoV-2 variant in Japan. (A) The light blue, dark blue, and green bars show the number of cases with unknown status of vaccine, the status of no vaccine and first dose of vaccine, and fully vaccinated status, respectively. (B) The light blue, dark blue, light green, and dark green bars show Alpha, Delta, Omicron, and other variants, respectively.

**Fig 2.**
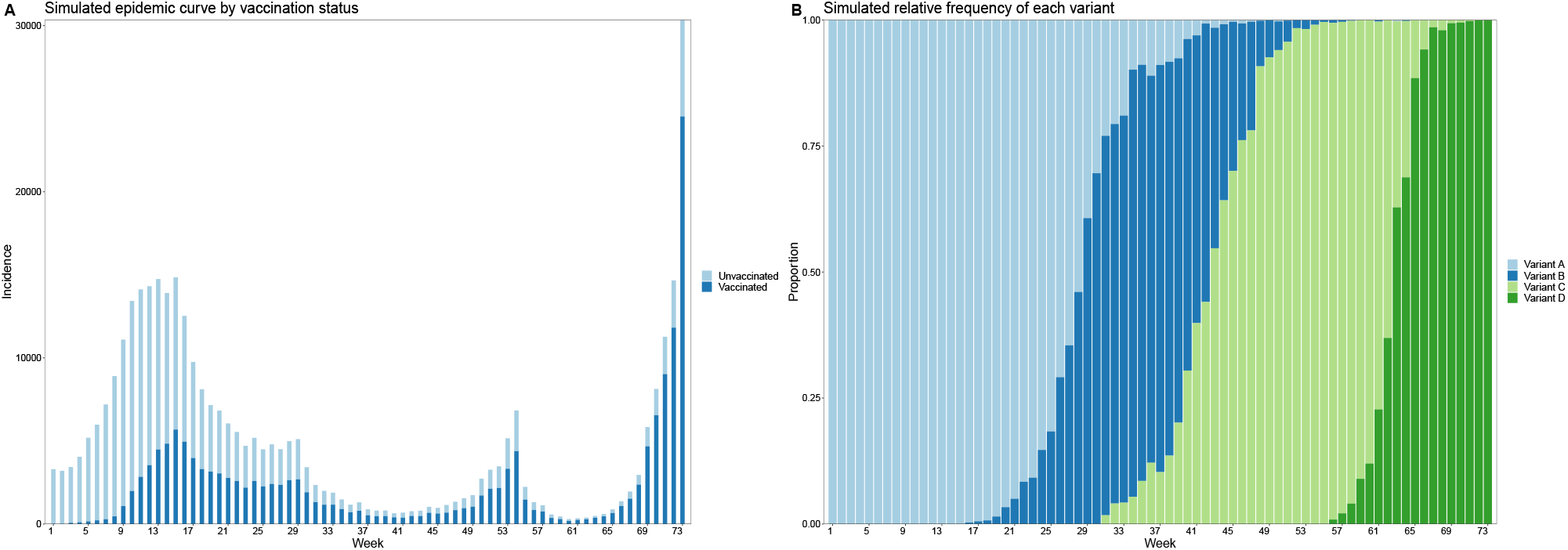
The simulated outbreak data in the presence of multi-variant circulation. (A) The simulated epidemic curve by vaccination status. (B) Cross-sectional proportion of the four variants in the simulation.

**Fig 3.**
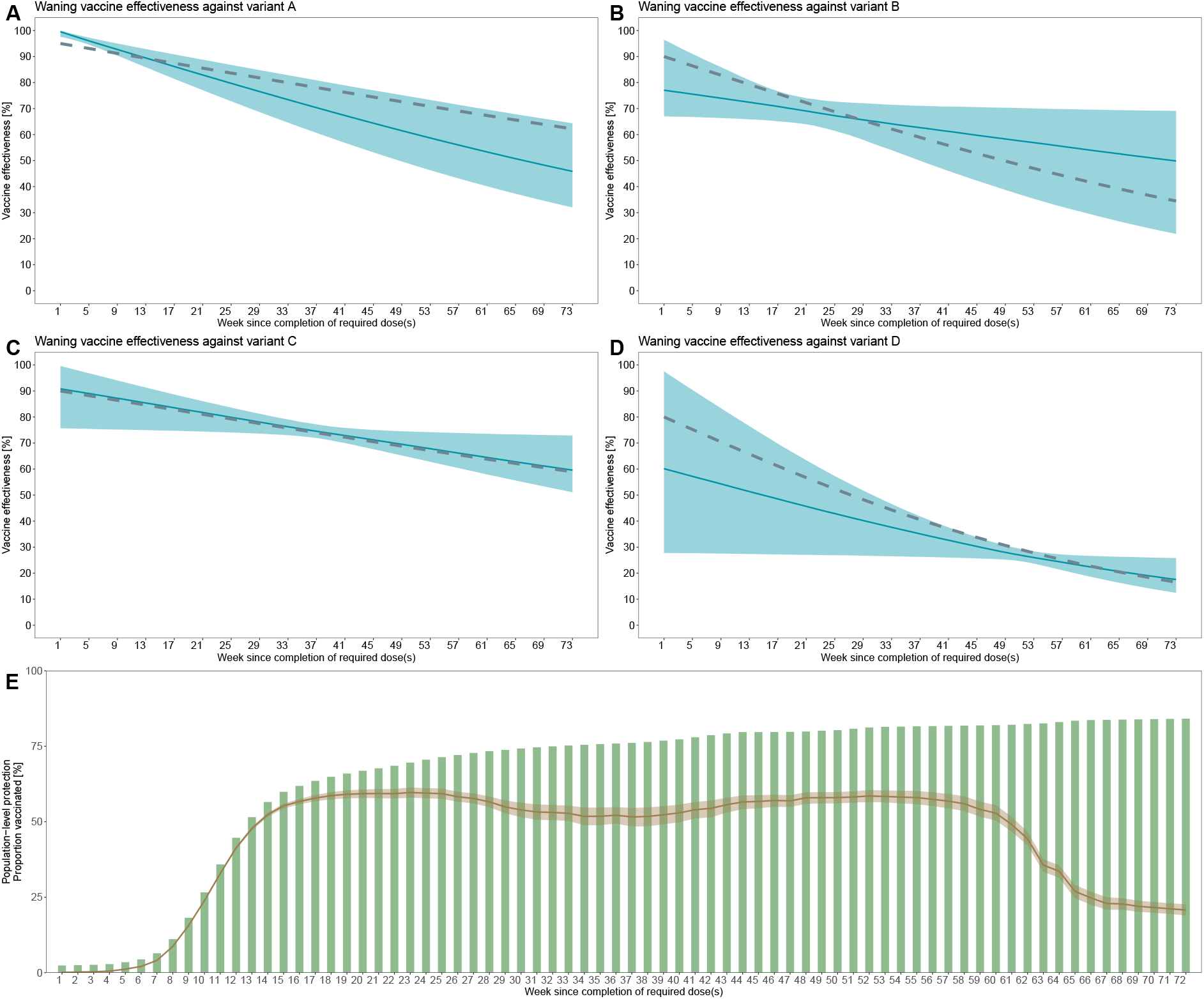
Estimated waning vaccine effectiveness with the exponential parametric model. (A)-(D) The estimated and ground truth variant-specific vaccine effectiveness. The light blue lines and shades indicate the medians and 95% credible intervals, respectively. The silver dotted lines are the ground truth vaccine effectiveness. (E) The cumulative proportion vaccinated (green bars) and the variant-weighted cross-sectional protection (median and 95% credible intervals denoted by brown lines and shades, respectively).

**Fig 4.**
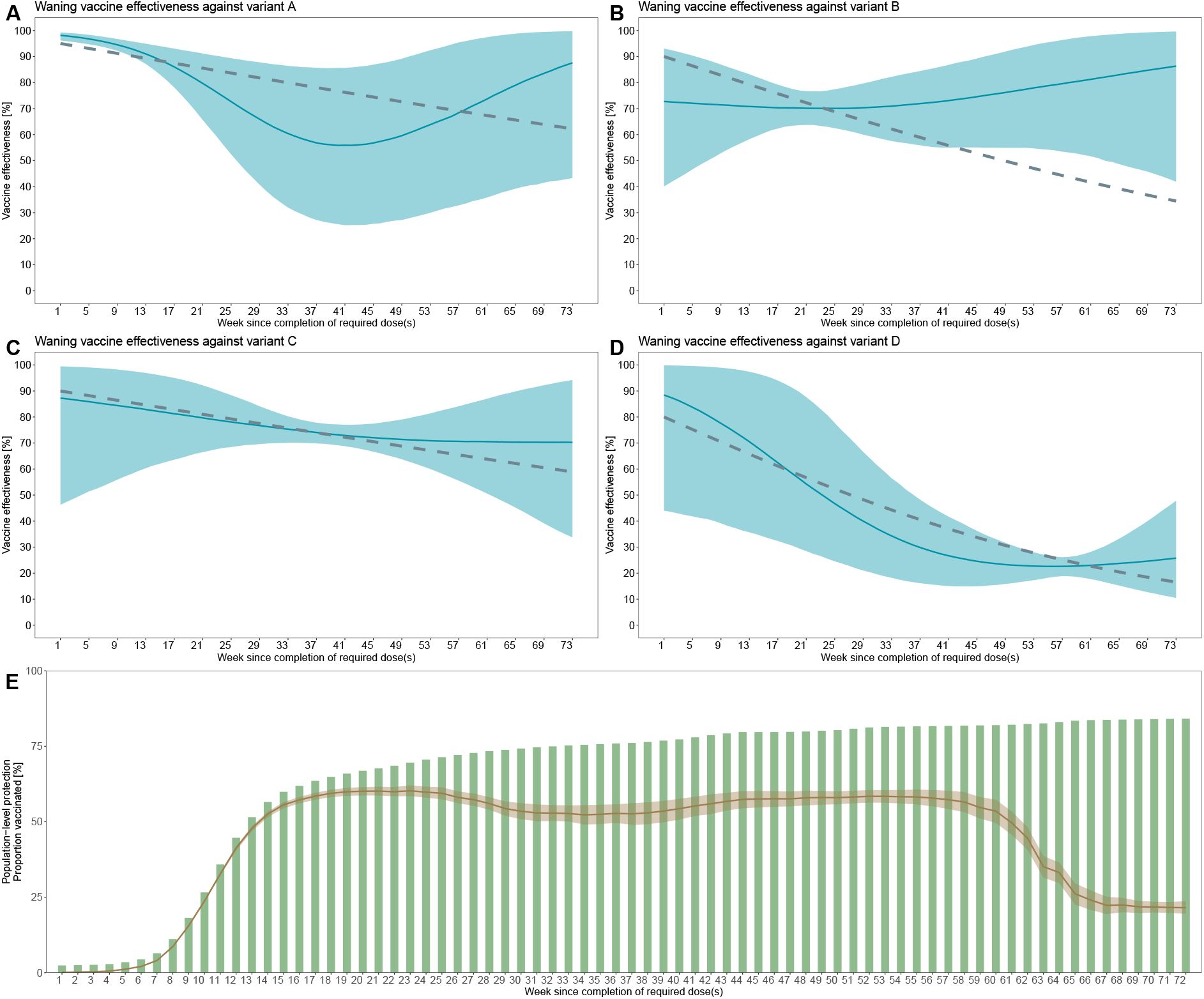
Estimated waning vaccine effectiveness with the semi-parametric model. (A)-(D) The estimated and ground truth variant-specific vaccine effectiveness. The light blue lines and shades indicate the medians and 95% credible intervals, respectively. The silver dotted lines are the ground truth vaccine effectiveness. (E) The cumulative proportion vaccinated (green bars) and the variant-weighted cross-sectional protection (median and 95% credible intervals denoted by brown lines and shades, respectively).

### Application to COVID-19 in Japan (2021–2022)

The estimated vaccine effectiveness against Delta and Omicron in the parametric approach with an exponential decay model is overall in a good accordance with the estimates from test-negative design studies using individual-level case records (Fig 5 (A) (B)). The estimated VCP showed a notable decline around the end of 2021 / beginning of 2022, which coincides with the emergence of the immune-escaping Omicron variant (Fig 5 (C)). The alternative parametric model (logistic curve) yielded essentially identical results (S1 Fig). Both of the parametric models proved a good fit to the data (S2 Fig, S3 Fig). The semi-parametric approach also yielded qualitatively similar results (Fig 6, Fig S4).

**Fig 5.**
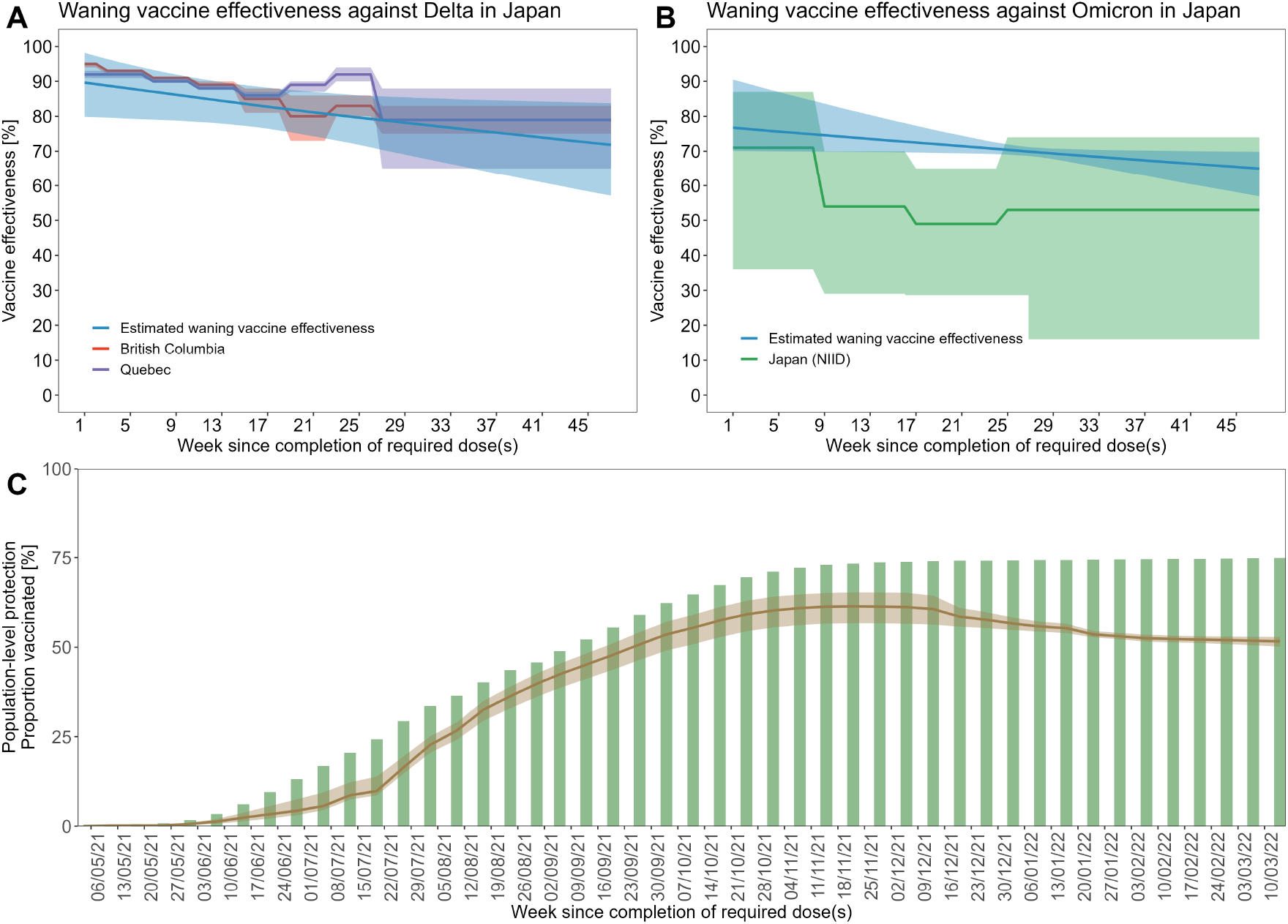
The waning of vaccine effectiveness estimated in parametric model with exponential function using Japan data. (A)(B) The estimated and ground truth variant-specific vaccine effectiveness. The blue lines and shades indicate the medians and 95% credible intervals, respectively. Reference values from other studies are also displayed with their 95% uncertainty bounds. (C) The cumulative proportion vaccinated (green bars) and the variant-weighted cross-sectional protection (median and 95% credible intervals denoted by brown lines and shades, respectively).

**Fig 6.**
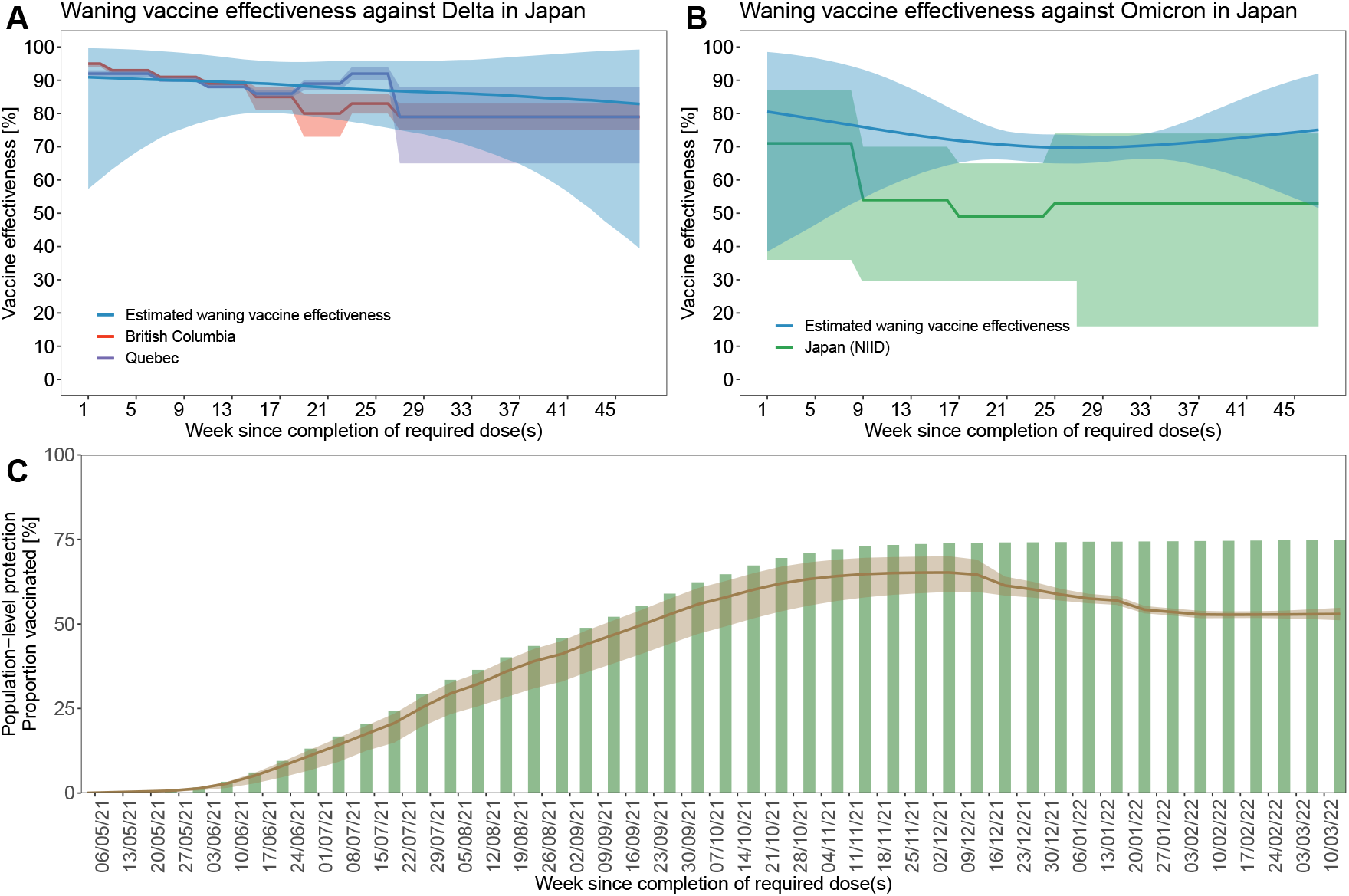
The waning of vaccine effectiveness estimated in semi-parametric model using Japan data. (A)(B) The estimated and ground truth variant-specific vaccine effectiveness. The blue lines and shades indicate the medians and 95% credible intervals, respectively. Reference values from other studies are also displayed with their 95% uncertainty bounds. (C) The cumulative proportion vaccinated (green bars) and the variant-weighted cross-sectional protection (median and 95% credible intervals denoted by brown lines and shades, respectively).

## Discussion

We have proposed a novel Bayesian framework that allows for crude but rapid estimation of variant-specific waning vaccine effectiveness using routinely-collected population-level surveillance data. Our application examples suggested that, with reasonable constraints on the functional form of waning, the proposed approach could plausibly produce estimates that are generally consistent with those from studies using individual-level case records. Moreover, the estimated VCP in our approach can be used to monitor the overall level of protection against the currently circulating variants in the population. Compared to those standard approaches to waning vaccine effectiveness that typically require extensive time and resource to construct fine-scale datasets, our approach could provide preliminary/complementary estimates within a shorter time frame, which would inform public health responses under rapidly evolving population immunity landscapes in the presence of multiple variants.

In our analysis, we used both the parametric (exponential or logistic curves) and semi-parametric approaches (cubic spline) to specifying the functional form of waning vaccine effectiveness. The parametric approaches could provide stable estimates if one can specify the model for the waning vaccine effectiveness whilst at the risk of bias and/or overconfidence if the model is misspecified. Semi-parametric approach is more robust to the risk of misspecification due to its flexibility, although it may become unreliable in the absence of sufficient data to inform the full range of the waning curve.

In our simulation, both approaches exhibited plausible performances overall, with the estimated waning curves for the co-circulating variants mostly in line with the ground truth. However, compared with the results in the parametric models (Fig 3, S1 Fig), the estimates from the semi-parametric model were partly less reliable or biased (Fig 4). This was likely because the simulated dataset did not include a sufficient number of cases that can inform those sections of the waning curve. The majority of vaccinated individuals were assumed to have had their dose around week 10–15 and they were: up to around 10 weeks since vaccination when variant A was dominant; and around 15–25 weeks, 30–50 weeks, and 50–65 weeks since vaccination when each of variant B, C, and D was dominant, respectively (Fig 2). These periods almost exactly correspond to the estimated waning curve for each variant was most certain and precise (similar patterns were also observed for the parametric approach; Fig 3). The waning curves for variant A and B showed a substantial deviation from the ground truth over the tail end of the curves. This is not unexpected because there was essentially no data corresponding to these sections; even the earliest-vaccinated individuals had not been past those many weeks since vaccination before these variants were almost extinct in the population. Meanwhile, waning curves for variant C and D did not show such a substantial deviation probably because the data existed for almost the entire sections (if scarce for some of them).

In contrast, the parametric model was able to capture the entire waning curves of variant-specific vaccine effectiveness because the model was less flexible and can extrapolate the sections for which the data was scarce. However, it should be noted that the success was largely due to the correctly specified functional form for waning. The results may be less reliable if the parametric model is misspecified. While it would be reasonable to impose some realistic constraints, e.g., monotonic decrease, on the functional form of waning curves to guide estimation, the possibility of misspecification should be carefully considered. Assessing concordance between the observed and modelled incidence (as shown in S2 Fig–S4 Fig) and comparing results from multiple candidate models (both parametric and semi-parametric) can be useful. In addition, as was the case for the semi-parametric model discussed above, one should also carefully assess which sections of the estimated waning curve was fully informed by the data.

In the application example to COVID-19 in Japan (Fig 5), estimated vaccine effectiveness against the Delta variant from both parametric and semi-parametric approaches were overall in line with those in existing test-negative design studies. The estimates of vaccine effectiveness against the Omicron variant were lower than those against the Delta variant, as had been suggested previously [27], although they lay slightly above the reference estimates from elsewhere [14]. The suggested difference in the waning curves between vaccine effectiveness against Delta and Omicron as well as the decline in the estimated VCP from the week of the emergence of the Omicron variant might have provided an early warning if our method had been applied to the surveillance data in a timely manner.

It should be noted that we made several simplifying assumptions in our study. First, we assumed that the contact behaviours of unvaccinated and vaccinated individuals were identical and that individuals from these groups mix nonassortatively (i.e. proportionate mixing [18–20]). Our estimation may be likely biased by heterogeneity that susceptibility differs amongst vaccinated individuals under not randomly mixing pattern [28]. Potential differences between the characteristics between unvaccinated and vaccinated individuals are typically addressed by adjustment for covariates in standard vaccine effectiveness studies where individual-level data is available. Such adjustment is inherently impossible in our approaches relying on population-level data, which constitutes one of the reasons why the results should be deemed preliminary. Second, we did not consider the depletion-of-susceptible bias, which typically arise when unvaccinated individuals are more likely to have experienced prior infection (thus to have developed immunity) than vaccinated individuals. Although this bias is suggested to be minor in the case of the current COVID-19 outbreak because of the high baseline vaccine effectiveness [29], the potential caveat needs to be recognised in a broader context (e.g., when applying our approach to other diseases). Third, our approach implicitly assumes that the epidemic is limited to a closed population (i.e., no imported cases), although the model could be extended to include imported cases if the distinction between imported and local cases is available in the dataset. Fourth, we excluded the effect of booster vaccines from analysis due to the limitation of the publicly available dataset. However, our framework could be naturally extended to jointly estimate waning vaccine effectiveness with primary and booster doses if incidence data and vaccine doses data are both separately available for booster vaccination.

Taken together, the present study provides a useful and practical tool to estimate waning vaccine effectiveness in multi-variant epidemics from population-level surveillance data. Continuous emergence of SARS-CoV-2 variants with different levels of immune-escape properties has highlighted the need for monitoring vaccine effectiveness as a dynamic metric shaped by waning and variant replacement. Our method would inform control efforts by providing the tentative yet timely estimates of vaccine effectiveness against co-circulating variants in the current and future pandemics.

## Data Availability

All the analysis was conducted in R v.4.0.2. Relevant data and code for the replication of this research is deposited on GitHub.

https://github.com/hiroaki-murayama/waning_ve_estimation_multivariant

## Supporting information

**S1 Fig. The waning of vaccine effectiveness estimated in parametric model with inverse logit function using Japan data**. (A)(B) The estimated and ground truth variant-specific vaccine effectiveness. The blue lines and shades indicate the medians and 95% credible intervals, respectively. Reference values from other studies are also displayed with their 95% uncertainty bounds. (C) The cumulative proportion vaccinated (green bars) and the variant-weighted cross-sectional protection (median and 95% credible intervals denoted by brown lines and shades, respectively)..

**S2 Fig. Simulation to check the fitness of the parametric model with exponential function to outbreak data in Japan**. The figures are the comparison between model-informed and observed epidemic curve with unvaccinated and vaccinated incidence. The dark blue bars show the observed incidence and the light blue bars show the estimated value of incidence out of posterior MCMC samples.

**S3 Fig. Simulation to check the fitness of the parametric model with logistic function to outbreak data in Japan**. The figures are the comparison between model-informed and observed epidemic curve with unvaccinated and vaccinated incidence. The dark blue bars show the observed incidence and the light blue bars show the estimated value of incidence out of posterior MCMC samples.

**S4 Fig. Simulation to check the fitness of the semi-parametric model to outbreak data in Japan**. The figures are the comparison between model-informed and observed epidemic curve with unvaccinated and vaccinated incidence. The dark blue bars show the observed incidence and the light blue bars show the estimated value of incidence out of posterior MCMC samples.

## Data availability

All the analysis was conducted in R v.4.0.2. Relevant data and code for the replication of this research is deposited on GitHub (https://github.com/hiroaki-murayama/waning_ve_estimation_multivariant).

## Author contributions

H.M. conceptualised the study. H.M., A.E., and S.Y. designed the study. H.M. implemented the models, collected and analysed the data, and wrote the first draft. All authors contributed to the writing and approved the final version of the manuscript.

## Acknowledgement

S.Y. and A.E. were supported by the Japan Society for the Promotion of Science (KAKENHI) under grant number 21K17713 (S.Y.) and 22K17329 (A.E.).

