## Supplementary for "Estimating waning vaccine effectiveness from population-level surveillance data in multi-variant epidemics"

### Supplementary Figure

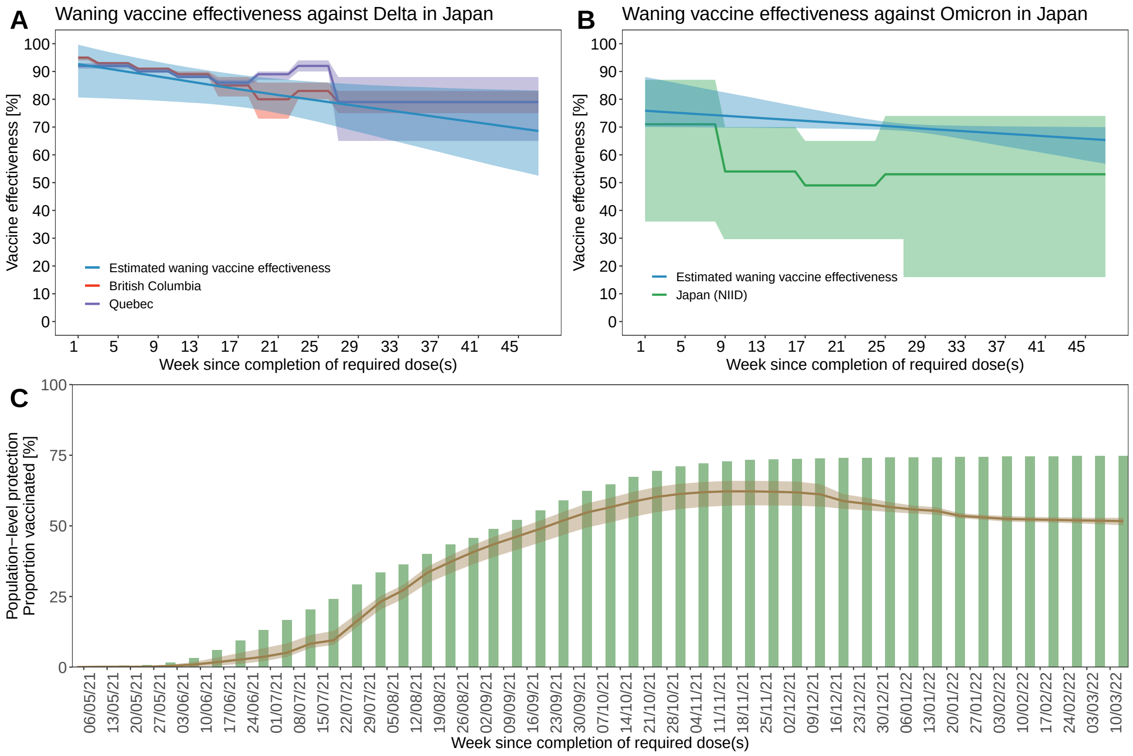


Figure S1 : The waning of vaccine effectiveness estimated in parametric model with inverse logit function using Japan data. (A)(B) The estimated and ground truth variant-specific vaccine effectiveness. The blue lines and shades indicate the medians and 95% credible intervals, respectively. Reference values from other studies are also displayed with their 95% uncertainty bounds. (C) The cumulative proportion vaccinated (green bars) and the variant-weighted cross-sectional protection (median and 95% credible intervals denoted by brown lines and shades, respectively).

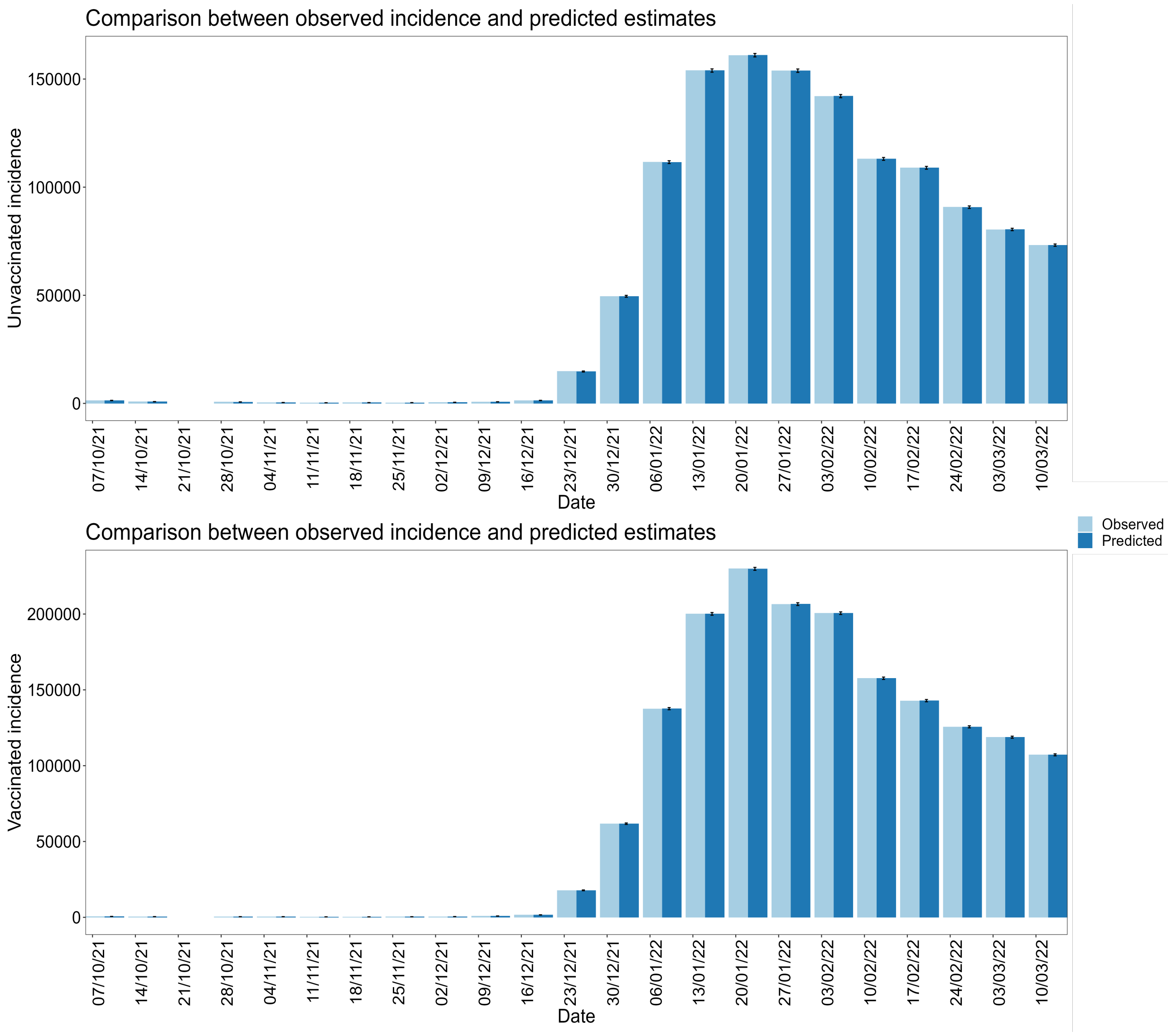


Figure S2 : Simulation to check the fitness of the parametric model with exponential function to outbreak data in Japan. The figures are the comparison between model-informed and observed epidemic curve with unvaccinated and vaccinated incidence. The dark blue bars show the observed incidence and the light blue bars show the estimated value of incidence out of posterior MCMC samples.

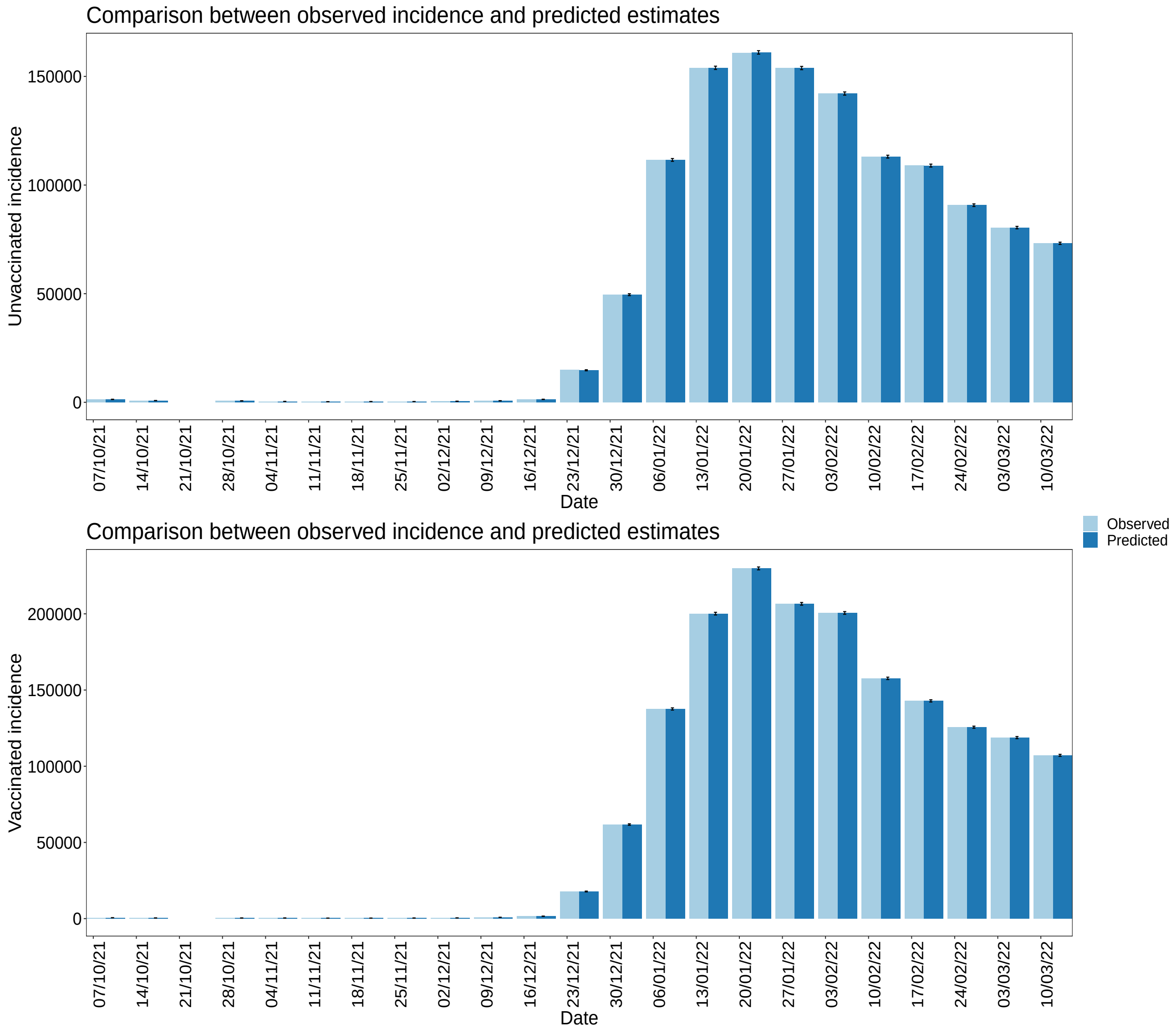


Figure S3 : Simulation to check the fitness of the parametric model with logistic function to outbreak data in Japan. The figures are the comparison between model-informed and observed epidemic curve with unvaccinated and vaccinated incidence. The dark blue bars show the observed incidence and the light blue bars show the estimated value of incidence out of posterior MCMC samples.

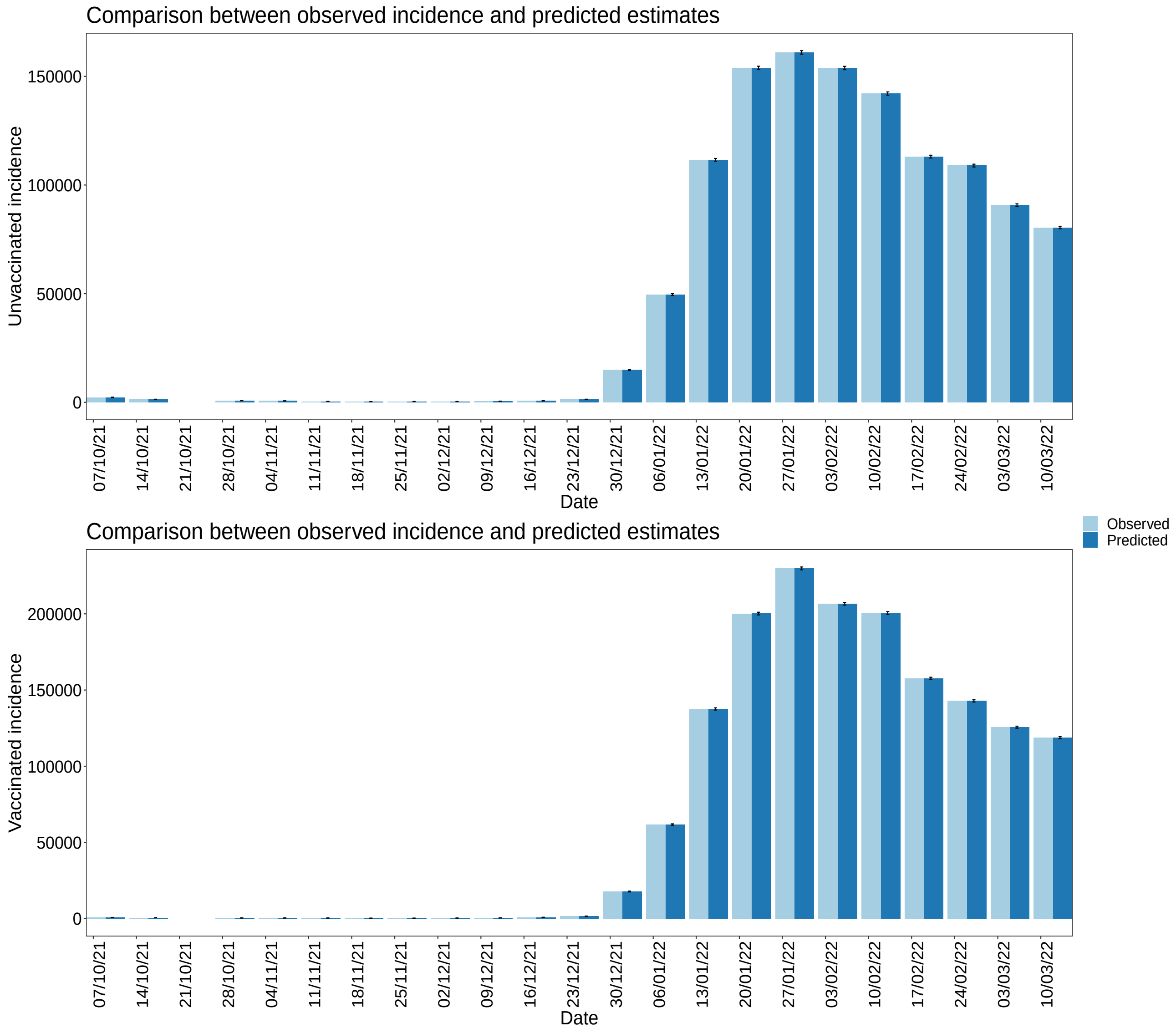


Figure S4 : Simulation to check the fitness of the semi-parametric model to outbreak data in Japan. The figures are the comparison between model-informed and observed epidemic curve with unvaccinated and vaccinated incidence. The dark blue bars show the observed incidence and the light blue bars show the estimated value of incidence out of posterior MCMC samples.
